# High Prevalence of Human and Cattle Fascioliasis in Zaria and Environs: An Emerging Zoonotic Problem

**DOI:** 10.1101/2025.11.15.25340295

**Authors:** Anthony Oche Ameh, Junaidu Kabir, Isa Danladi Jatau, Saaondo James Ashar, Ahmad Bello Kaoje, Lauratu Lawal Halilu, Olanrewaju Eyitayo Igah, Usman Isa, Beatty-Viv Maikai

**Affiliations:** Ahmadu Bello University, Zaria, Kaduna State, Nigeria; National Veterinary Research Institute, Vom; Ministry of Agriculture, Kaduna

**Keywords:** Fascioliasis, *Fasciola*, Humans, Animals, Cattle, Bile, Cross-sectional study, Prevalence, Enzyme-Linked Immunosorbent Assay, Risk factors

## Abstract

**Background:** Fascioliasis is an important global neglected tropical foodborne zoonosis. It causes economic losses in animals and recurrent health challenges in humans. It is misdiagnosed with other conditions which present similar signs, as a result, its true prevalence is not known. This study reports the current prevalence of fascioliasis and the association between fascioliasis and risk factors in cattle and humans in Zaria and environs, Kaduna, Nigeria.

**Methods:** This was a cross-sectional study involving 202 slaughter cattle and 186 human subjects. Formol ether sedimentation and enzyme linked immunosorbent assay techniques were used for sample examination. Chi-square was used to test for association between *Fasciola* species infection and factors like age, sex, breed and body condition score. Kappa statistic (K) was used to determine the level of agreement between the tests used.

**Results:** The prevalence of Fasciola species infection in cattle was 76 (37.6%) and humans was 183 (98.4%). Formol ether and bile sedimentation techniques had slight agreement (K = 0.041) in detecting Fasciola species infection in cattle. Fasciola species infection was highest in female human subjects 102 (54.8%) and adults (25 years and 59 years) 77 (41.6%). Fascioliasis was not significantly associated with breed, age and body condition score in cattle as well as gender and age in human subjects.

**Conclusion:** Fascioliasis is highly prevalent in humans and cattle indicating its emergence and endemicity respectively in the study area and the need for intervention.

## Introduction

Fascioliasis is an important global foodborne parasitic helminth disease affecting livestock and man and is classified amongst the list of neglected tropical diseases by the World Health Organization (WHO) [1, 2]. It is caused by a parasitic trematode of the genus *Fasciola* commonly referred to as liver flukes of the family *Fasciolidae. Fasciola hepatica* and *Fasciola gigantica* are the most economically important species [3].

Fascioliasis is present across all continents except Antarctica and amongst the diseases caused by helminths, it has the widest distribution longitudinally, latitudinally and altitudinally. *Fasciola hepatica* predominates Europe and America, but both species overlap in distribution in many parts of Africa and Asia [4]. *Fasciola gigantica* has a more constricted geographical distribution occurring mostly in Africa, the Middle East and Asia [4].

The lifecycle of *Fasciola* involves a final host (ruminants) and an intermediate host (a mollusc) for its larva stage development [5]. The definitive hosts acquire infection after ingesting the infective metacercarial stage when grazing or in drinking water [6]. Humans are infected with *Fasciola* from consumption of vegetables contaminated with infective metacercaria [7].

In cattle, fascioliasis causes substantial economic impact affecting 10% to 80% of beef and milk producing cattle in the developing and developed nations [8]. The resultant losses arise from a decreased milk yield, reduced weight gain, poor quality of carcass, cost of treatment and control, death and condemnation of affected livers across abattoirs in Nigeria [9]. Losses due to liver condemnation resulting from fascioliasis alone, amounted to about US$134,000 in South-western Nigeria [10].

On the other hand, human fascioliasis (HF) was regarded to be of less significance till 1990, when it became important following progressive reports and studies of human endemic regions coupled with increased reports of human cases [11]. HF is highly pathogenic causing immune suppression in its migratory, invasive and biliary stages of infection, affecting most inhabitants of endemic and hyperendemic regions [11, 12]. Due to its related abdominal complications, HF is estimated to result in 90,000 Disability Adjusted Life Years (DALYs) affecting about 17 million people globally [13]. In extreme cases, it results in highly pathogenic conditions as ophthalmic and neurologic syndromes bringing about lasting sequalae and is indicative of its public health significance [12].

An upsurge in the incidence rate of this emerging disease is expected due to factors like: climate change, livestock trade, human migration, globalization, and modifications of the environment, all of which will enhance the parasite transmission and abundance of the intermediate host [14] which will remain undetected a cnd unreported if not investigated, hence the need to carry out this epidemiologic investigation.

Although with respect to prevalence and intensities, it is thought that the expected correlation between animal and human fascioliasis only appears at a basic level, but infection rates may differ significantly in human endemic regions while in regions where animal fascioliasis occur, the prevalence of human fascioliasis may appear insignificant [15, 16]. This may not necessarily be the case as fascioliasis is not routinely screened in healthcare facilities particularly in Nigeria, and may be misdiagnosed with other diseases which present similar symptoms of fever and malaise like malaria. Therefore, there is need for epidemiologic investigation to describe the prevalence and distribution of fascioliasis in animals and humans. This study therefore aimed to investigate the prevalence of fascioliasis in cattle and humans, determine the association between fascioliasis and associated factors in cattle and humans.

## Methods

### Study Location

This study was conducted in Zaria and its environs which encompass Giwa, Sabon Gari and Zaria Local Government Areas (LGAs). Zaria is located within latitudes 11° 7̍ꞌ, 11° 12ꞌ N and longitudes 07° 41ꞌE in the Northern Guinea Savannah zone. The population of Zaria as estimated by the National Population Commission (NPC) comprises a total of 698,348 inhabitants with 406,990 inhabitants in Zaria and 291,358 inhabitants in Sabon gari LGAs [17]. Next to Kaduna, the state capital, Zaria is the second largest urban centre in Kaduna state. The urban centre encompasses Zaria and Sabon Gari LGAs. Hausa and Fulani tribes inhabit the present day old-walled Zaria town. There have been alterations of the natural vegetation in this Guinea Savannah zone due to different human activities like deforestation, construction and overgrazing [18].

Zaria is characterised by rain-fed agricultural activity spanning from May to October and irrigation farming in the dry season between November and April. It has a tropical climate with mean and annual rainfall of 1092.8mm and a monthly mean temperature which ranges between 13.8°C to 36.7°C [19].

Zaria and environs, has three main slaughter facilities for cattle and three major general hospitals in each of the three LGAS which are accessed by the residents and were used for this study. The slaughter facilities are: Zaria Main Abattoir; Zaria and Giwa slaughter facilities located in Sabon gari, Zaria and Giwa LGAs respectively. The general hospitals are: Major Ibrahim Bello Abdullahi (MIBA); Hajiya Gambo Sawaba (HGS) and General Hospital Giwa located in Sabon gari, Zaria and Giwa LGAs respectively.

Agriculture is the main source of livelihood for approximately 40-75% of the working population in these LGAs and cattle in these LGAs are mostly extensively managed and this is usually marked by low productivity, diseases, hazards and predisposition to potential infection sources for *Fasciola* species [20]. The slaughter facilities generate improperly managed waste and effluents which impact the environment [21].

### Study Population

This was a cross-sectional study involving cattle and human populations. The samples from the cattle were obtained from three slaughterhouses for a period of three months. Human samples were obtained from human subjects attending 3 general hospitals for a period of three months. The human subjects came to the hospitals for other tests or routine check-up.

### Sample Size Determination

A prevalence (p) of 13.67% obtained from a previous study by Liba *et al*. [22] and 3.5% from a study by Isah and Dalhatu [23] were used to determine the sample size for cattle and humans respectively. The sample sizes were estimated using the formula by Thrusfield [24]. At a confidence interval of 95%, with n being the required sample size; d the margin of error (5%) and z being the z-score (1.96) obtained from the normal distribution, the sample size was estimated to be 202 samples for cattle and 62 samples for human subject by employing a 10% and 20% non-response respectively.

### Sampling procedure

The cattle and human subjects to be sampled were selected using systematic random sampling. The number of cattle selected from each slaughter facility was proportional to the total number of cattle slaughtered at each facility. Faecal and bile samples were collected from each selected cattle. On the other hand, at each general hospital, for spread of sample, 62 subjects were selected. Faecal and blood (for serum) samples were obtained from each selected subject.

### Ethical Consideration

Ethical clearance for this study was approved and obtained from the Kaduna State Ministry of Health Ethical Review Committee with number NHREC/17/03/2018. Consent forms in English and Hausa languages containing a comprehensive explanation of the aim, possible risk and benefits of the study were administered to the participants of this study. Only the consent form signed by either willing subjects or parents or legal guardians for subjects less than 18 years were recruited for this study. The study posed minimal risk to the subjects and all data collected and results were kept confidential. Links were only made to subjects for the purposes of treatment or management. All subjects positive for *Fasciola* species were referred to their consulting physician for further management.

### Sample Collection

About 10g of faeces was collected directly from the rectum of selected cattle using a clean polythene bag worn over a gloved hand. The faecal samples were placed in wide mouth screw top containers and labelled properly. Approximately 10 ml of bile was obtained directly from the gall bladder into wide mouth screw top containers and were labelled immediately.

The human subjects were given a wide mouth screw top container to bring about 5 g of their faecal sample. The collection of blood samples from human subjects was done by a phlebotomist by obtaining 3mls of blood via venipuncture of the basilic or median cubital vein according to the method described by CLSI [25]. The blood was then transferred immediately into plain labelled sample 10 ml bottles. The blood sample was then centrifuged at 2,000 rpm for two minutes in order to obtain serum. The serum was then carefully extracted using sterile Pasteur pipettes and deposited in clean properly labelled 5 ml serum vials and stored at – 25 ° C in a deep freezer for subsequent processing. All samples were transported on ice in plastic containers to the Parasitic Zoonoses Laboratory, Department of Veterinary Public Health and Preventive Medicine, Ahmadu Bello University, Zaria for subsequent processing and examination.

### Sample Processing and Examination

Formol ether sedimentation technique as described by Arora and Brij [26] was used for the detection of *Fasciola* eggs. The processed faecal samples were examined under the microscope at 10× magnification for *Fasciola* eggs characterized by its large, oval and yellowish-brown colour with a distinct flat operculum [27].

Biles samples were processed by dispensing about 4ml of the bile sample into a labelled test tube followed by addition of 1ml of 10% formalin. The mixture was allowed to stand for 5 minutes, after which 1ml of diethyl-ether was added. The ensuing mixture was corked in the test tube and vigorously shaken to ensure proper mixing, and then centrifuged for 10 minutes at 2000 rpm. After centrifugation, the supernatant was decanted, however a few drops were left with the sediment. The sediment was dispensed using a pipette on a clean glass slide with a cover slip placed over it and examined under the microscope at 10× magnification for *Fasciola* eggs characterized by its large oval and yellowish-brown colour with a distinct flat operculum [27].

The sera samples were analyzed using AccuDiag™ *Fasciola* IgG Enzyme Linked Immunosorbent Assay [ELISA] Kit from Diagnostics Automation/Cortez Diagnostics Incorporated USA according to the directions of the manufacturer.

### Data Analysis

Statistical Package for Social Science (SPSS) version 25 was used for data analysis. Chi-square was used to determine association between *Fasciola* species infection and factors such as age, sex, breed of animals. Pearson chi-square and Fishers exact tests were also used to determine the association between *Fasciola* species infection and gender and age in human subjects where appropriate. All P-values less than or equal to 0.05 (P ≤ 0.05) were considered to be statistically significant.

Kappa statistic (K) was used to determine the agreement between faecal and bile sedimentation techniques for detecting *Fasciola* infection in cattle. Kappa statistic was also used to determine the agreement between faecal sedimentation and serology in the detection of *Fasciola* infection in human subjects. The results of the Kappa test were interpreted according to the scale by Landis and Koch [28] as follows: < 0: Poor agreement; 0.0 – 0.20 indicating slight agreement; 0.21 – 0.40 indicating fair agreement; 0.41 – 0.60 indicating moderate agreement; 0.61 – 0.80 indicating substantial agreement and 0.81 – 1.00 indicating almost perfect agreement.

The age of human subjects was disaggregated into 7 groups as described by Diaz *et al.* [29] into: young children (1-4 years), older children (5-9 years), young adolescents (10-14 years), older adolescents (15-19 years), young adults (20-24 years), adults (25-59 years), older adults (60 years and above).

## Results

*Fasciola* species detection by bile sedimentation 76 (37.6%) in cattle and Enzyme Linked Immunosorbent Assay (ELISA) 183 (98.4%) in humans were high. Furthermore, there was slight agreement (K = 0.041) between formol ether and bile sedimentation techniques as well as formol ether sedimentation and ELISA (K= 0.004) in the detection of *Fasciola* species infection in cattle and human subjects respectively (Table 1).

**Table 1:** Prevalence of *Fasciola* species infection in Cattle and Humans in Zaria and environs, Kaduna State, Nigeria.

|  |  |  |  |
| --- | --- | --- | --- |
|  | **Total number examined** | **Number positive (%)** | **Kappa Statistic (K)** |
| **Cattle** |  |  |  |
| Formol ether sedimentation | 202 | 30 (14.9) | 0.041<sup>a</sup> |
| Bile sedimentation | 202 | 76 (37.6) |  |
| **Humans** |  |  |  |
| Formol ether sedimentation | 179 | 17 (9.5) | 0.004<sup>b</sup> |
| ELISA | 186 | 183 (98.4) |  |
| Formol ether sedimentation and ELISA | 179 | 17 (9.5) <sup>c</sup> |  |
| The superscript ‘a’ indicates slight agreement between faecal and bile sedimentation techniques in the detection of *Fasciola* species infection in cattle.<br>The superscript ‘b’ indicates slight agreement between faecal sedimentation and ELISA techniques in the detection of *Fasciola* species infection in human subjects.<br>The superscript ‘c’ indicates human subjects positive by both tests |  |  |  |

Subsequent test for association using Pearson Chi-square (χ^2^) for possible risk factors and fascioliasis, showed that there was no statistically significant association between breed (χ^2^ = 2.255, P = 0.521); age (χ^2^ = 0.117, P = 0.733) and body condition score (χ^2^ = 0.605, P = 0.739) and fascioliasis in cattle in the study area (Table 2).

**Table 2:**
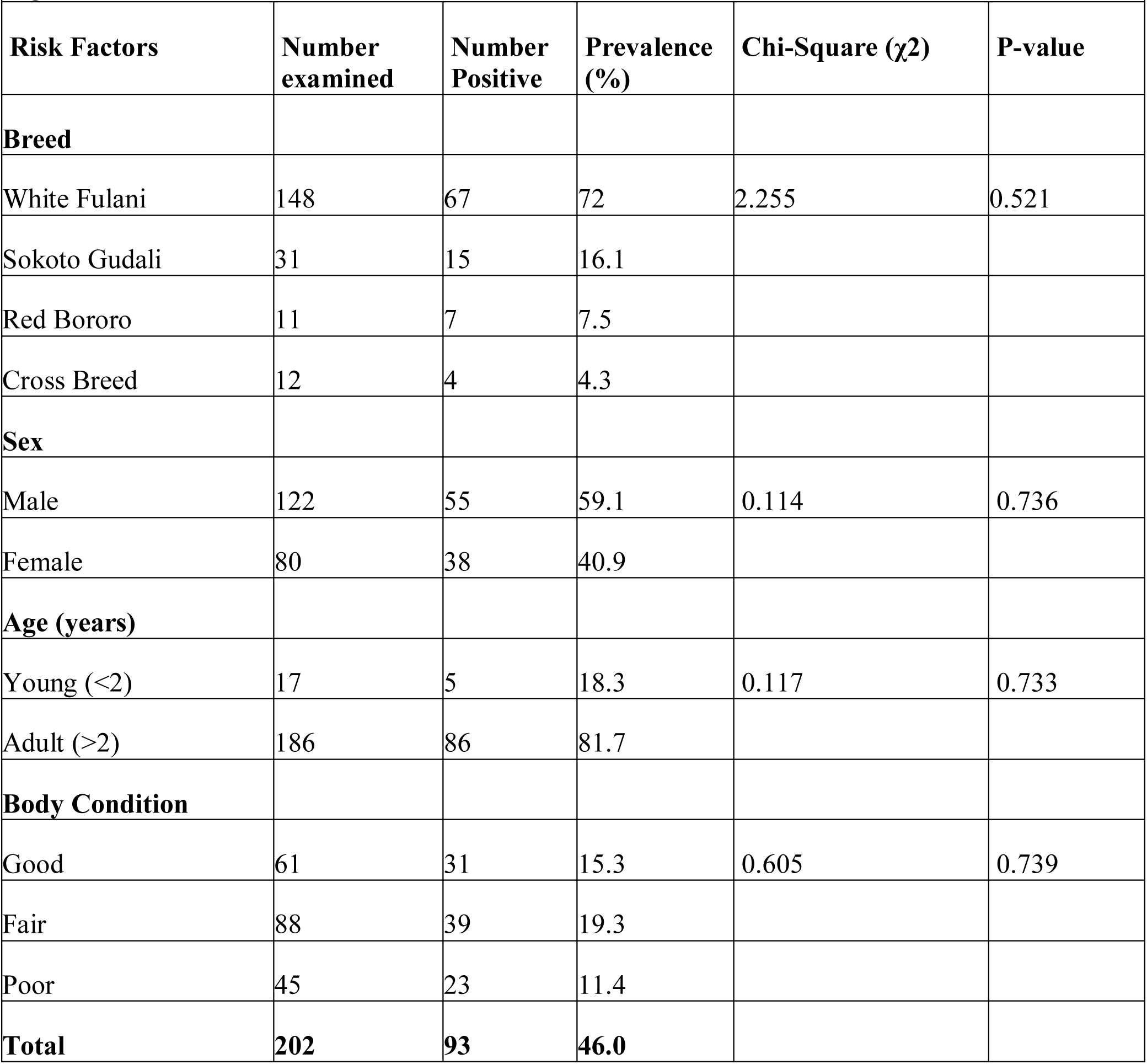
*Fasciola* species infection based on risk factors in cattle in Zaria and environs, Kaduna State, Nigeria.

Also, *Fasciola* species infection was higher in female subjects 102 (54.8%) than in male subjects 81 (43.5%), with no statistically significant association (χ^2^ = 0.600, P = 0.439) observed (Table 3). Based on age, *Fasciola* species was most prevalent in adults aged between 25 years and 59 years 77 (41.6%) with no statistical association (Fisher’s exact value = 5.439, P = 0.536) observed.

**Table 3:** *Fasciola* species infection in relation to risk factors in Human subjects in Zaria and environs, Kaduna State, Nigeria.

|  |  |  |  |  |  |
| --- | --- | --- | --- | --- | --- |
| **Risk Factors** | **Number examined** | **Number Positive** | **Prevalence (%)** | **Chi-Square ($\chi^2$)** | **P-value** |
| **Sex** |  |  |  |  |  |
| Male | 83 | 81 | 43.5 | 0.600 | 0.439 |
| Female | 103 | 102 | 54.8 |  |  |
| **Age (years)** |  |  |  |  |  |
| 1-4 (young children) | 4 | 4 | 2.2 | 5.439 | 0.536 |
| 5-9 (older children) | 9 | 9 | 4.9 |  |  |
| 10-14 (young adolescents) | 13 | 12 | 6.5 |  |  |
| 15-19 (older adolescents) | 51 | 50 | 27.0 |  |  |
| 20-24 (young adults) | 23 | 23 | 12.4 |  |  |
| 25-59 (adults) | 78 | 77 | 41.6 |  |  |
| 60 and above (older adults) | 8 | 8 | 4.3 |  |  |
| **Total** | **186** | **183** | **98.4** |  |  |

## Discussion

This study further confirms the endemic and emerging nature of fascioliasis in Nigeria with evidence of on-going silent infection in cattle and humans in Zaria and environs, Kaduna State. Previously, fascioliasis was considered a disease primarily of livestock, however, it is now regarded as an important emerging zoonotic disease affecting man [14]. The high prevalence of fascioliasis in cattle from this study while similar to those reported by Ieren *et al*., [30]; Dada and Jegede [31] may be indicative of the endemic nature of this disease in cattle. Consequently, this may have accounted for the high prevalence of the disease in the human populace thereby corroborating findings by Mas-Coma et al. [16], that the presence and operation of abattoirs lacking basic hygiene, cold chain distribution, and biosafety facilities contribute to increased transmission of fascioliasis in humans as found in this study.

Additionally, the high prevalence of fascioliasis in cattle and human subjects may be linked to drivers of zoonotic infection particularly, environmental modifications (irrigation), livestock and human migration coupled with increased livestock trade and economic activities [32, 33]. Also, Mas-Coma et al. [16] reported a high likelihood of transmission of fascioliasis from animal to humans, particularly where there is close interaction between animals and humans.

Some of these drivers are obtainable in the study area where majority of cattle ownership as well as those slaughtered at abattoirs were brought from the North-western and North-eastern parts of Nigeria where the grazing system is that of the nomadic system by the Fulani herdsmen. This nomadic system usually involves the practice of moving cattle in search of pastures and watering points across different parts of Nigeria, thus exposing them to potential sources of infection as they ingest pastures with infective metacercariae. Consequently, these cattle are slaughtered across abattoirs in the study area which generate improperly managed waste materials [21]. These waste materials such as faeces and other effluents constitute the main environmental challenge as they contain large quantities of solid and liquid wastes which subsequently affect ground water quality when discharged into water channels and streams located away from the abattoir [34]. Also, these wastewater effluents, contaminate water sources for domestic use or are used as irrigation sources for growing salad vegetables like lettuce which can lead to transmission of zoonotic infections like fascioliasis [35] when such vegetables are consumed without maintaining proper washing and hygiene. These vegetables which may contain infective metacercaria are usually harvested fresh and eaten raw as salad thereby affecting the populace in the study area. As a result, these exposures could result in increased infectivity and the high prevalence in cattle and humans recorded in the study area.

Fascioliasis is reported to be emerging and re-emerging in African, Asian and Middle eastern countries [36, 37]. However, with respect to human fascioliasis, some countries still lack data despite advances in techniques for surveillance and diagnosis while there is abundance of data supporting animal fascioliasis in others. The challenges associated with the diagnosis of fascioliasis in humans were previously linked to the different phases of infection, migration capacities, heterogenous clinical nature and immunological intricacies of the parasite. Nonetheless, the basic diagnostic techniques for diagnosis of human fascioliasis have in recent times been advanced. It was resolved that no single diagnostic method encompasses all the different previously raised factors. Hence, a combination of different techniques including at least a blood and stool technique was recommended [12] Therefore, this study utilized a combination of blood and stool techniques. This may have likely accounted for the high detection by ELISA, 183 [98.4%] and formol ether sedimentation 17 (9.5%) in blood and stool respectively.

Thu et al. [38] recorded a similar seroprevalence (95.1%) in Vietnam and the finding from this study also corroborates validation studies by Tran *et al*. [39]. Additionally, all the 17 human subjects positive for *Fasciola* species eggs by formol ether sedimentation were also positive by ELISA screening. Among the tests for detecting *Fasciola* species infection, immunodiagnostic tests are very useful with respect to epidemiological collection, clinical diagnosis and specific antibodies identification with great accuracy [40]. The gold standard for human *Fasciola* species infection diagnosis is faecal egg detection, but the rate of detection of ova is quite low because during the acute and migratory phase of infection eggs may not be laid by the worms. Also, in cases of ectopic fascioliasis, the eggs may not be detectable and in chronic infections, there is an intermittent shedding of eggs. The recommended confirmation of fascioliasis according to the World Health Organization (WHO) is by clinical signs together with either parasitological or immunological methods [41].

Furthermore, a major benefit of the antibody detection in fascioliasis is that close to two weeks after infection, antibodies are detectable unlike ova detection in faeces 2 to 3 months after infection. Such early detection and proper therapy may prevent damage to the hepatic tissues. Hence, for prompt detection and therapy, antibody detection is a suitable technique [42].

The high serological prevalence for *Fasciola* species infection recorded in this study may be because serological tests especially for Immunoglobulin G (IgG) may not differentiate current or active infections from previous infections [43]. Also, antibodies may persist for a minimum of 4 to 5 months and in some cases, years after treatment. Notwithstanding, Apt *et al*. [44] reported absence of *Fasciola* antibodies two months post treatment while Tawfeek and Hussein [45] reported a negative IgG result using ELISA in 80% and 95% of cases within the first and after four months respectively.

Also, the prevalence of 17 (9.5%) by formol ether technique for human subjects recorded in this study was higher than that reported in other parts of Nigeria, like Kano (0.9%) by Ihesiulor *et al*. [46] and Bauchi (3.5%) by Isah and Dalhatu [23]. This may likely be due to the physiological status of the sample populations in both studies. This study focused on human subjects attending general hospitals while the others dealt with apparently healthy populations like school children, thereby accounting for the higher prevalence recorded in this study.

The physiological condition or state of animals is an important contributory risk factor associated with helminthosis which affects productivity, thereby resulting in economic losses [47]. However, in this study, breed, sex, age and body condition score, were not significantly associated with *Fasciola* species infection. Other compounding factors like the management system of rearing cattle, the nutritional status of the cattle amongst others may have resulted in infection observed in this study. These findings agreed with those of Elelu *et al*. [48]; Eze and Briggs [49]. However, Okoh et al. [50] and Liba et al. [22] found significant association between *Fasciola* species infection and sex, attributing possible causes to disparities in intrinsic factors (immunology, physiologic and genetic composition of both sexes) and extrinsic factors (environmental conditions of rearing).

Relatedly, no significant association between *Fasciola* species infection and gender was recorded in human subjects. This finding is similar to a study by Esteban *et al*. [51] in Egypt. Similarly, a study in the Bolivian Altiplano by Parkinson *et al*. [52] demonstrated that there was no significant association between *Fasciola* species infection and gender in a study involving 8000 subjects. Differences in gender with respect to helminth infection are rarely observed in food borne parasitic infection, hence, interventions should cut across all genders.

*Fasciola* species infection among various age groups showed no statistical significance. This contrasts studies by Cabada *et al.* [53], who reported independence between the risk factor for liver fluke infection and an increased age especially in children (11 years on average). This result further contrasts a case series of *Fasciola* species infection reported by de Van *et al*. [54] in toddlers and infants in Vietnam. A total of 5 children were reported to have had acute and chronic infections with an additional 38 cases detected in children below 4 years between 1856 and 2016 [54].

The relationship between *Fasciola* species infection and the age in children has not been well demonstrated in this study and region. However, it has been shown in endemic regions that children assist their parents in farming activities and animal rearing, making them spend many hours away from home. Consequently, they eat or suck many wild vegetables which exposes them to infective metaceracariae [55]. Dietary habits such as eating leafy vegetables particularly those grown close to grazing fields; drinking contaminated and untreated water enhance the risk of *Fasciola* species infection [47].

## Limitations

Sampling of human subjects was limited to only those willing to consent and the use of only IgG ELISA kit may not have differentiated current or active infections from previous infections. However, this was the first study investigating human fascioliasis in the study area.

## Conclusion

There is high prevalence of fascioliasis in cattle (37.6%) further confirming its endemicity in the study area. Also, the high serological detection of fascioliasis in humans (IgG = 98.4%) in the study area indicates it is an emerging zoonosis. Fascioliasis affects all genders and age groups in humans in the study area. Also, fasciolosis affects all breeds, sexes and age groups of cattle in the study area

## Recommendations

There should be public health enlightenment campaigns, periodic mass screening and deworming programmes targeting animals and humans in the study area. Also, cattle faeces from abattoirs should not be used for manure without proper treatment (composting) while abattoir waste and effluents should be properly disposed of or channelled into pits to prevent their runoff and possible contamination of water sources for irrigation and consumption. These measures will break the developmental and transmission cycle of the parasite in the study area.

### What is already known on this topic

- Fascioliasis is an important global foodborne Neglected Tropical parasitic helminth disease affecting livestock and man.
- Fascioliasis is emerging and re-emerging in African, Asian and Middle eastern countries.
- Fascioliasis causes significant economic losses in animals and is highly pathogenic in humans

### What this study adds

- This is the first serological evidence of human fascioliasis in the study area and is indicative of its emerging zoonotic nature.
- This study also revealed that *Fasciola* infection was not related to or dependent on the age or gender of human subjects in the study area, indicating that there is an equal opportunity for infection across all groups, therefore, interventions should target all groups in the study population.
- This study further confirms the endemicity of fascioliasis in cattle in the study area and as potential reservoirs for zoonotic infection.

## Data Availability

all data produced in the present work are contained in the manuscript

## Acknowledgements

The authors appreciate the staff of Zaria main abattoir, Giwa and Zaria city slaughter facilities, Department of Veterinary Public Health and Preventive Medicine, Ahmadu Bello University, Zaria as well as those of Major Ibrahim Bello Abdullahi Hospital; Hajiya Gambo Sawaba Hospital and General Hospital Giwa for their assistance in the conduct of this study.

## Authors’ contributions

Conceptualization and design of the study: Junaidu Kabir, Isa Danladi Jatau, Beatty-Viv Maikai, Anthony Oche Ameh.

Supervision: Junaidu Kabir, Isa Danladi Jatau, Beatty-Viv Maikai.

Data collection and curation: Anthony Oche Ameh, Saaondo James Ashar, Ahmad Bello Kaoje, Lauratu Lawal Halilu, Usman Isa.

Formal Analysis: Junaidu Kabir and Anthony Oche Ameh.

Manuscript Draft and revision: Junaidu Kabir, Anthony Oche Ameh, Isa Danladi Jatau, Olanrewaju Eyitayo Igah, Saaondo James Ashar, Ahmad Bello Kaoje, Lauratu Lawal Halilu, Usman Isa.

